# A tale of three SARS-CoV-2 variants with independently acquired P681H mutations in New York State

**DOI:** 10.1101/2021.03.10.21253285

**Authors:** Erica Lasek-Nesselquist, Janice Pata, Erasmus Schneider, Kirsten St. George

## Abstract

Several SARS-CoV-2 variants of concern have independently acquired some of the same Spike protein mutations - notably E484K, N501Y, S477N, and K417T - associated with increased viral transmission and/or reduced sensitivity to neutralization by antibodies. Repeated evolution of the same mutations, particularly in variants that are now rapidly spreading in various regions of the world, suggests a fitness advantage. Mutations at position P681 in Spike – possibly affecting viral transmission - have also evolved multiple times, including in two variants of concern. Here, we describe three variants circulating in New York State that have independently acquired a P681H mutation and the different trajectories they have taken. While one variant rose to high prevalence since later summer 2020 it appears to be in decline. The other two variants were more recently detected in New York and harbor additional Spike mutations that might be cause for continued monitoring. The latter two P681H variants have shown moderate increases in prevalence but ultimately all might be subject to the same fate as more competitive variants come to dominate the scene.

## Introduction

New York State has served as a bellwether during the SARS-CoV-2 pandemic in the United States (US). It was the initial epicenter of the pandemic in the US, highlighting both the effects of delayed containment efforts and successful mitigation strategies (Rosenberg et al. 2020; Lasek-Nesselquist, Singh, et al. 2021; Maurano et al. 2020; Gonzalez-Reiche et al. 2020). An outbreak in Westchester County led to the widespread distribution of the 20C clade, demonstrating the rapidity with which SARS-CoV-2 could spread through a community and beyond (Lasek-Nesselquist, Singh, et al. 2021; Deng et al. 2020; Zhang et al. 2020; Gonzalez-Reiche et al. 2020). New York was also one of the first to identify the presence of the B.1.1.7 variant in the US (associated with 40-70% increased growth rate; (Volz et al. 2021) and possibly 30% increased mortality rate; Iacobucci 2021), serving as both a source and a sink for this variant of concern (Alpert et al. 2021). Recently, the number of B.1.526 variants harboring Spike E484K or S477N mutations (possibly enabling increased transmission and/or evasion of monoclonal antibodies) were identified as rapidly growing within the state (Annavajhala et al. 2021; Lasek-Nesselquist, Lapierre, et al. 2021; West et al. 2021), underscoring the need for fortified surveillance efforts.

Here we report on the trajectory of three SARS-CoV-2 variants in New York with P681H mutations in the Spike protein. The Spike protein is required for entry into the host cell and is composed of an S1 subunit that attaches to the host cell receptor angiotensin-converting enzyme 2 (ACE2), an S2 subunit necessary for fusion between virus and cell membranes, and a multibasic furin cleavage site that separates S1 and S2 subunits upon cleavage, thereby initiating subsequent events that lead to membrane fusion (Shang et al. 2020; Hoffmann, Kleine-Weber, Schroeder, et al. 2020; Walls et al. 2020; Bosch et al. 2003). P681 is adjacent to the furin cleavage site, which facilitates efficient SARS-CoV-2 transmission and infection (Peacock et al. 2020; Hoffmann, Kleine-Weber & Pöhlmann 2020). Mutations at this location (P681H and P681R) have been acquired by the B.1.1.7 variant rapidly spreading around the world and the A.23.1 lineage, recently reported as the dominant lineage in Uganda (Rambaut et al. 2020; Bugembe et al. 2021). Currently, the P681H mutation is the most commonly identified Spike mutation in SARS CoV-2 genomes from New York State (aside from D614G, which characterizes the B.1. lineage and is essentially ubiquitous in the US). While the majority of these genomes are members of the B.1.243 lineage, we detected independently acquired P681H mutations in several lineages belonging to clades 19B, 20A, 20C, 20D, and 20G. Notably, we identified a lineage B.1.1.222 variant (clade 20B) with both P681H and T478K Spike mutations and a lineage B.1 variant (clade 20C) variant with Spike mutations P681H and S494P. T478 and S494 sit within the receptor binding domain (RBD) responsible for binding the spike protein to the ACE2-receptor of host cells. Mutations at both positions were shown to reduce the activity of some monoclonal antibodies (Liu et al. 2021; Greaney et al. 2021a; Weisblum et al. 2020). Evidence of convergent evolution as well as the structural location of P681 suggest that mutations at this position could contribute to an adaptive advantage. While the two variants described here show only moderate increases since their first appearances in New York, the combination of P681H and T478K or P681H and S494P mutations could confer additional benefits to the virus and are worthy of continued surveillance. New York is thus an experimental field, where variants endowed with differing fitness compete. It remains to be seen whether the P681H variants described here gain a foothold or succumb to more dominant forms on the rise.

## Results

As of 2020-03-04, over 12% of all SARS-CoV-2 genomes sequenced in New York carried a P681H mutation (Figure 1). After an initial peak from September to November 2020, the proportion of genomes with this mutation remained stable until a small increase in early winter 2020 (Figure 1). The rapid spread of B.1.1.7, which also has a P681H mutation as well as outbreaks in congregant settings, likely contributed to the increase in later months (Figure 1). In fact, excluding B.1.1.7 genomes from the analysis revealed a small decline in the number of P681H-containing genomes by the end of February 2021 (Figure 1).

**Figure 1A).**
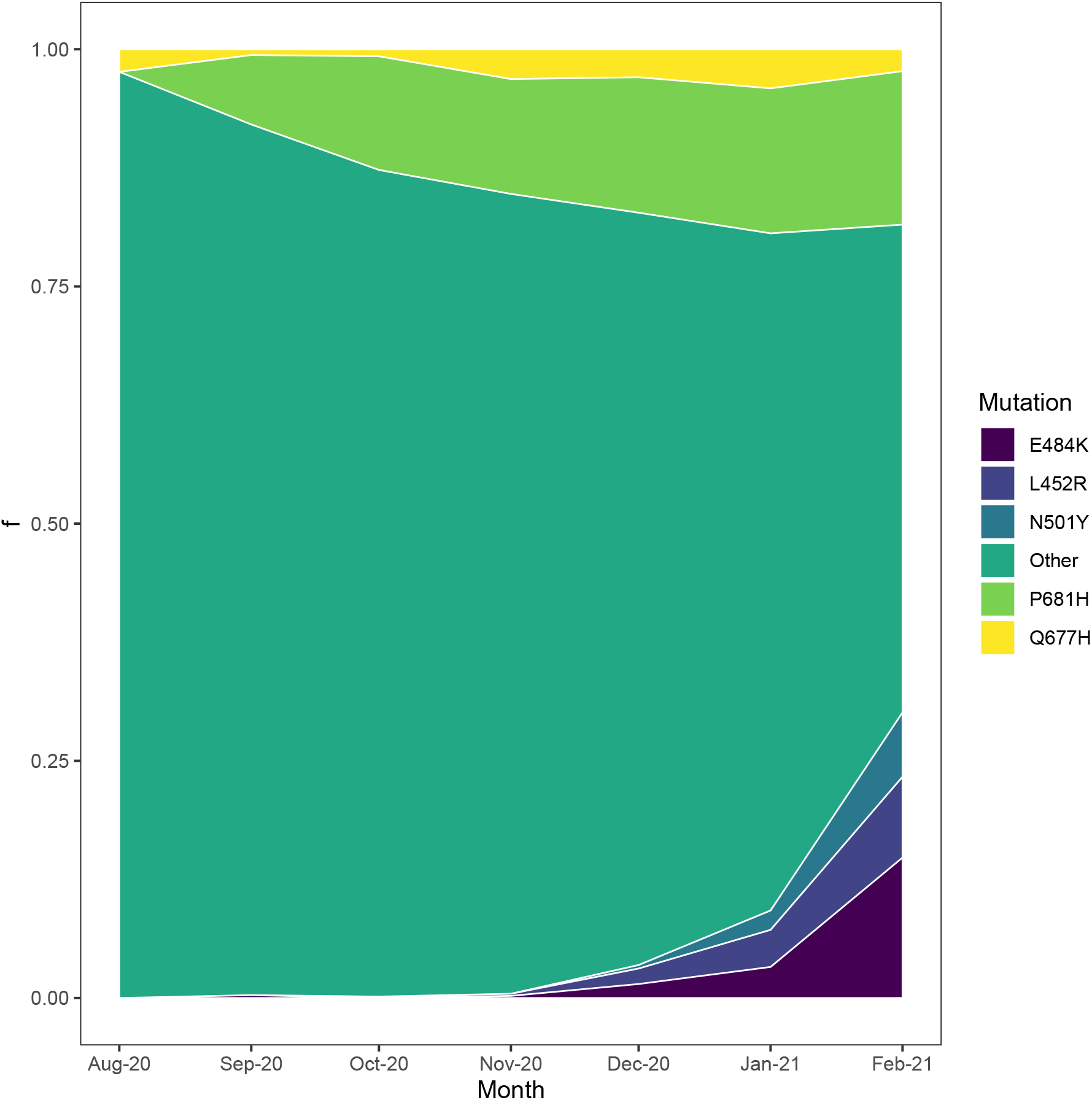
Proportion of genomes containing mutations of interest in New York State. Stacked area plot of several mutations detected in SARS-CoV-2 genomes from New York. E484K is associated with three variants of concern: P.1, B.1.351, and B.1.526. L452R is characteristic of B.1.429 and B.1.427 lineages, which were identified as variants of concern in California. N501Y is also present in three variants of concern: B.1.1.7, B.1.351, and P.1. Other is the frequency of all other spike mutations. Q677H has recently been noted for its independent evolution in multiple lineages (Hodcroft et al. 2021). P681H currently represents the most frequently encountered spike mutation (aside from D614G) in SARS-CoV-2 genomes from New York.

**Figure 1B).**
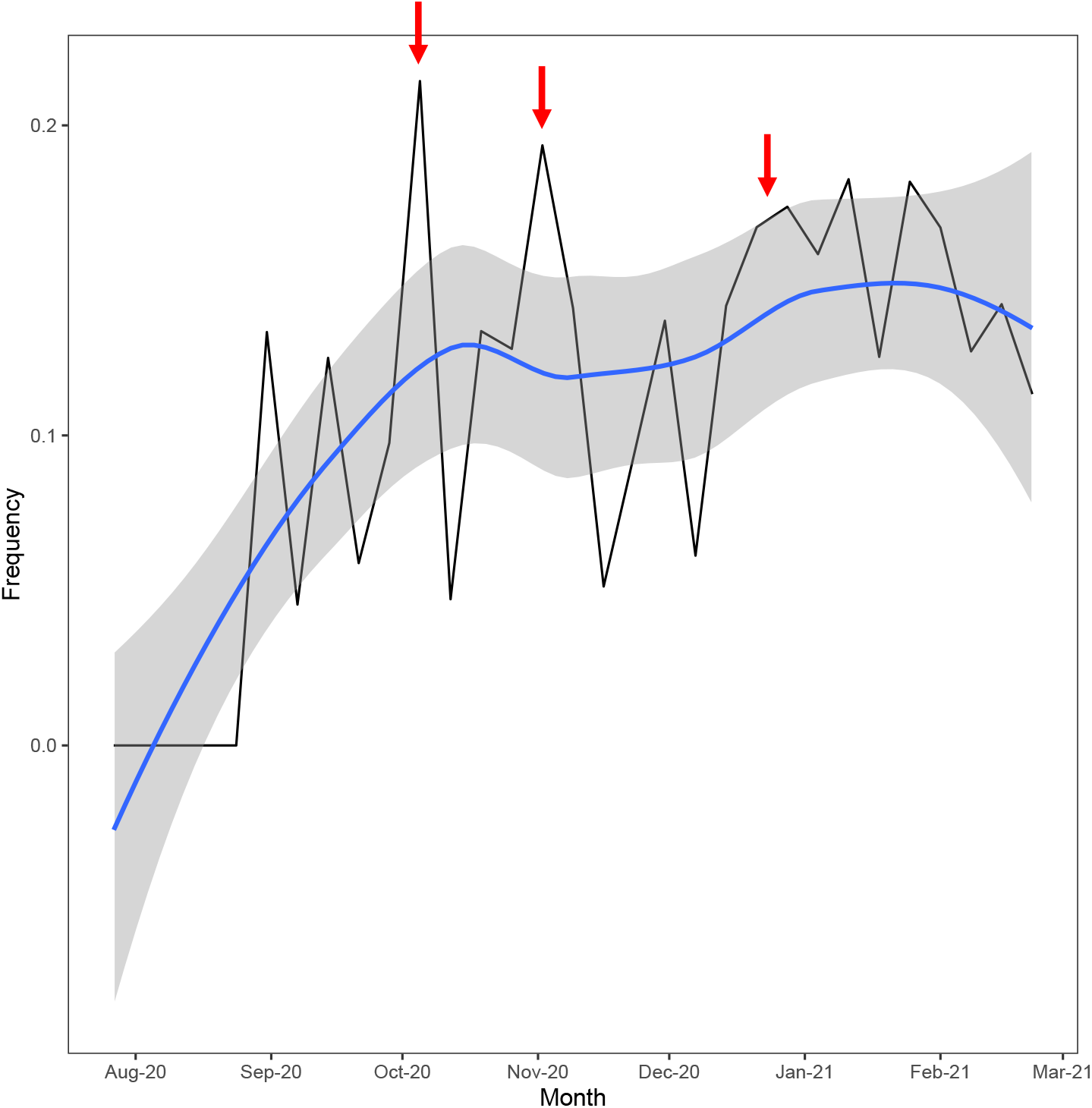
Frequency of P681H variants in New York as a function of time. The frequency of genomes with a P681H mutation in New York, excluding B.1.1.7 variants, calculated on a weekly basis (black line). The regression line (blue) was computed by the loess method. The gray shaded area represents the standard error or confidence intervals associated with the regression line. Red arrows indicate approximate timing of outbreaks due to B.1.243 variants in congregant settings.

The majority of SARS-CoV-2 genomes within New York state that carry a P681H mutation are members of the B.1.243 lineage (clade 20A; 63%; Supplementary Figure 1), which is detected mostly within the United States (https://cov-lineages.org/lineages). Phylogeographic analyses indicated that B.1.243 likely emerged in California mid-March 2020 (CA 100% CI, 2020-03-10 to 2020-03-20) with the P681H mutation acquired shortly thereafter (Figure 2). The B.1.243 variant was probably introduced in early August 2020 to New York (New York 100% CI, 2020-07-20 to 2020-08-22; Figure 2).

**Figure 2.**
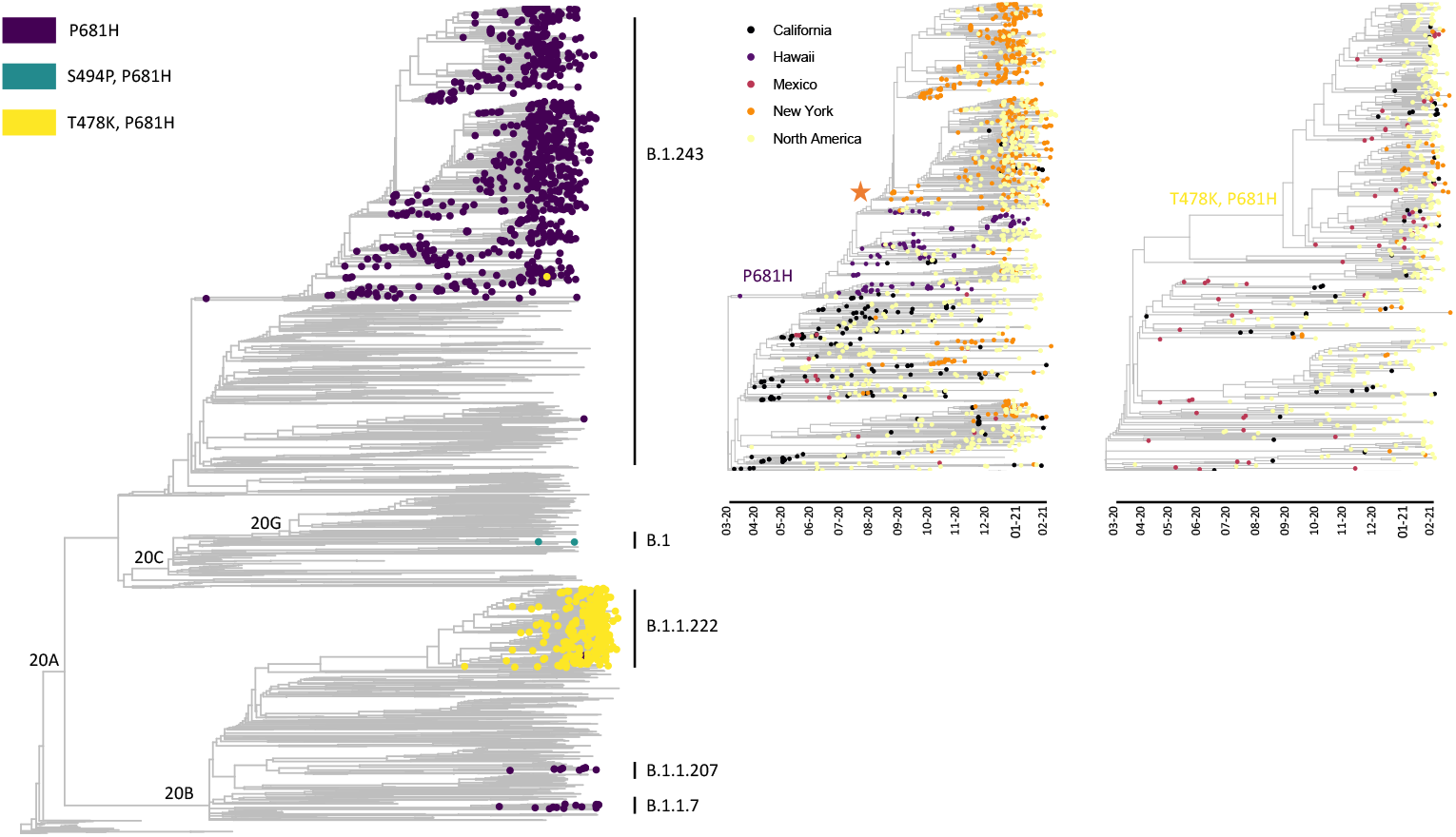
Phylogeny of SARS-CoV-2 highlighting the emergence of P681H in multiple lineages circulating in New York. Phylogeny of SARS-CoV-2 genomes sampled from New York and globally to highlight the independent evolution of P681H mutations in multiple clades and lineages (left panel). Time resolved subtrees showing the acquisition of the P681H mutation in the B.1.243 lineage (middle panel) and the T478K and P681H mutations in B.1 (right panel). The orange star indicates the estimated introduction of the B.1.243 variant into New York. Introductions of B.1.243 and B.1.1.222 from California and Mexico, respectively, are supported by the locations of basal genomes (middle and left panels).

Aside from California, New York and Hawaii have sequenced the greatest proportion of B.1.243 genomes in the US and have the largest number of B.1.243 genomes with a P681H mutation (Figure 3). While the B.1.243 variant appears to have mostly displaced the parental form in New York, other locations with both forms do not demonstrate the same trend (Figure 3). Similarly, the proportion of SARS-CoV-2 genomes assigned to the B.1.243 lineage with a P681H mutation varies dramatically by state and sampling effort (Supplementary Table 1). For example, California has sequenced ∼0.5% of its COVID-19 cases, of which only 0.5% of those sequenced are the B.1.243 variant (Supplementary Table 1). Washington sequenced ∼2.5% of all COVID-19 cases but the percentage of B.1.243 variants remained the same as that of California (Supplementary Table 1). In contrast, Hawaii has sequenced ∼3.4% of its COVID-19 cases and 76% are characterized as B.1.243 with a P681H mutation (Supplementary Table 1). Further, the proportion of B.1.243 variants in New York has dramatically declined as others, such as B.1.1.7 and B.1.526, have entered the arena (Figure 4). Thus, the foothold this variant had gained in certain locations, mainly Hawaii and New York, could reflect the effects of random processes, such as founder effects, rather than an adaptive advantage.

**Figure 3.**
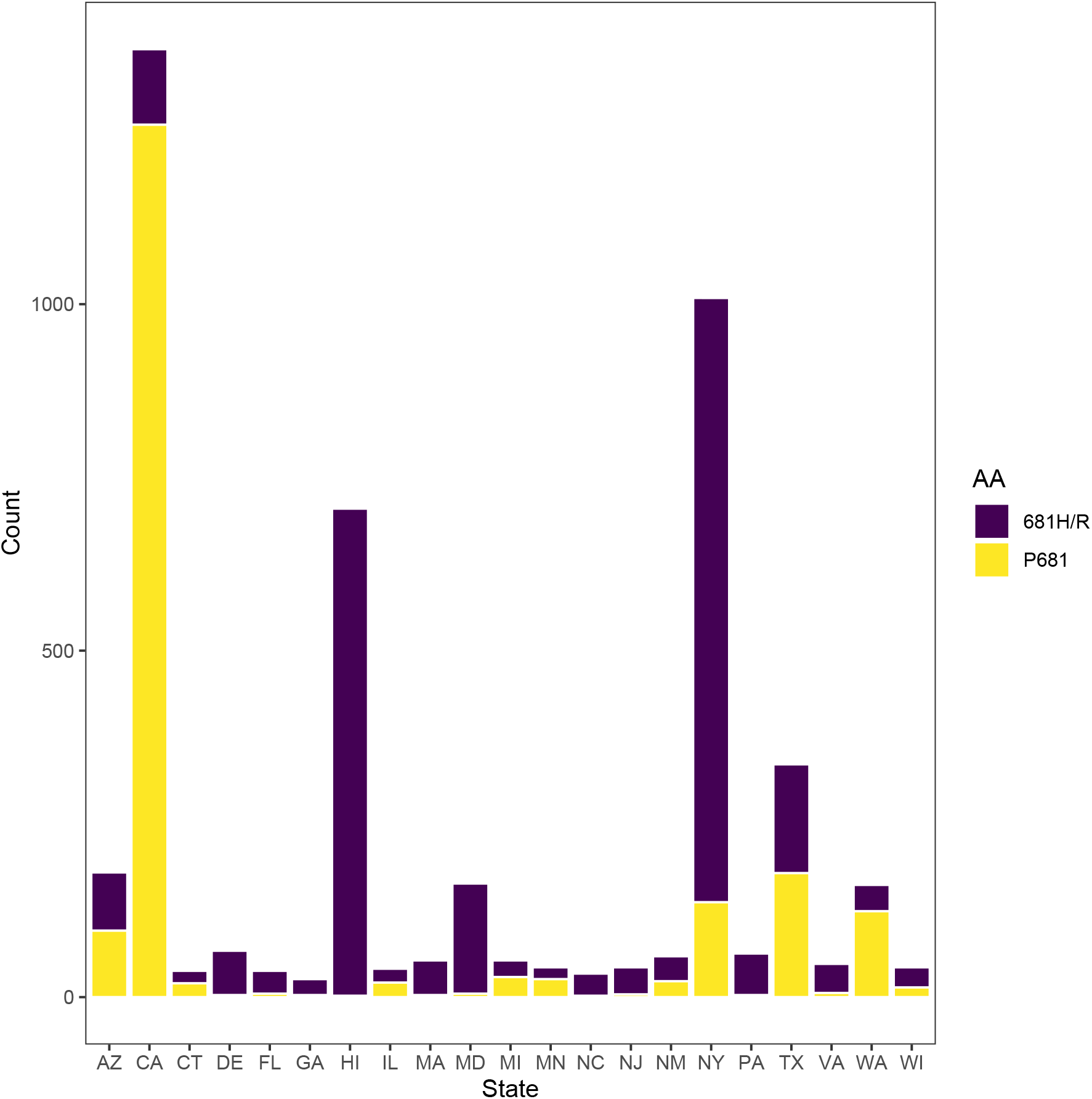
Number of B.1.243 genomes containing P681 or 681H amino acids for select states. Stacked barplot showing the number of B.1.243 SARS-CoV-2 genomes with and without P681H mutations sequenced for 21 states. The 21 states selected had the greatest proportion B.1.243 genomes with P681H mutations (as of 2021-02-26).

**Figure 4.**
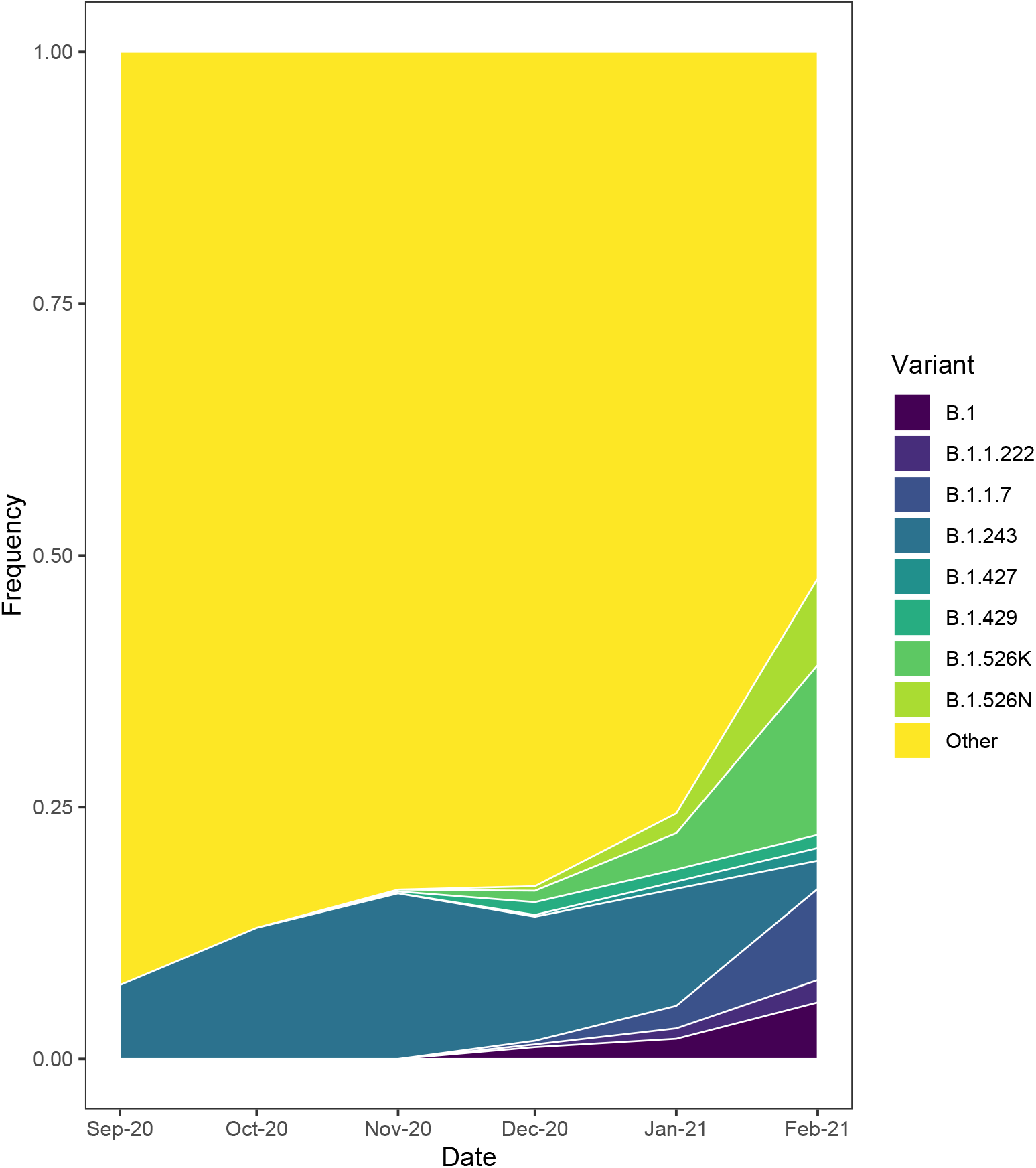
Stacked area map of SARS-CoV-2 variants in New York. The change in frequency of select SARS CoV-2 variants in New York as a function of time. B.1, variant with P681H and S494P mutations; B.1.1.222, variant with T478K and P681H mutations; B.1.1.7 the highly transmissible variant first identified in the United Kingdom (Rambaut et al. 2020); B.1.243, variants with a P681H mutation; B.1.427 and B.1.428 variants with an L452R mutation that are rapidly spreading in California (Zhang et al. 2021; Tchesnokova et al. 2021); B.1.526K and B.1.526N, B.1.526 variants with an E484K or S477N mutation rapidly spreading in New York (Lasek-Nesselquist, Lapierre, et al. 2021; Annavajhala et al. 2021; West et al. 2021); Other, all other lineages detected in New York.

A second variant carrying a P681H mutation has emerged within the 20B clade, lineage B.1.1.222, which also harbors a T478K mutation in the spike protein. Phylogeographic analyses suggest that the B.1.1.222 variant emerged in Mexico around Mid-September 2020 (Mexico 100% CI, 2020-08-26 to 2020-09-29; Figure 2) with subsequent spread, including to 48/50 states (the greatest proportion occurring in Texas, California, and New York; Supplementary Table 2). The B.1.1.222 variant first appeared in New York on 2020-12-23 and now represents the majority of B.1.1.222 genomes (66%; Table 2) from multiple introductions into the state (Figure 2). However, B.1.1.222 variants have increased only ∼2% since December 2020 and represent a minor fraction of the circulating viral population in New York (Table 1 and Figure 4).

**Table 1.** Number and proportion of variants in New York for December 2020 to February 2021. Count, the total number of variants for each month; Percent, the proportion of the total number of SARS-CoV-2 genomes sequenced (n = 2197, 3079, 2612 for December, January, and February, respectively as of 2021-03-04) that the variants represent; Increase, the percent difference between February 2021 and December 2020.

| **Variant** | **Dec-20** |  | **Jan-21** |  | **Feb-21** |  | **Increase** |
| --- | --- | --- | --- | --- | --- | --- | --- |
|  | **Count** | **Percent** | **Count** | **Percent** | **Count** | **Percent** |  |
| B.1.526 484K | 25 | 1.14 | 111 | 3.61 | 441 | 16.88 | 15.75 |
| B.1.526 477N | 10 | 0.46 | 61 | 1.98 | 225 | 8.61 | 8.16 |
| B.1.1.7 | 8 | 0.36 | 69 | 2.24 | 215 | 8.23 | 7.87 |
| **B.1** | **26** | **1.18** | **62** | **2.01** | **147** | **5.63** | **4.44** |
| **B.1.1.222** | **6** | **0.27** | **32** | **1.04** | **58** | **2.22** | **1.95** |
| B.1.427 | 4 | 0.18 | 22 | 0.71 | 33 | 1.26 | 1.08 |
| B.1.429 | 28 | 1.27 | 37 | 1.20 | 34 | 1.30 | 0.03 |

A third variant carrying a P681H mutation evolved independently within the 20C clade, lineage B.1, with an inferred date of introduction into New York in early November 2020 (NY 100% CI 2020-10-05 to 2020-12-06; data not shown). Additionally, this variant harbors T716I and S494P mutations in the spike protein, the latter of which has been shown to reduce the efficacy of some monoclonal antibodies (Greaney et al. 2021; Liu et al. 2021; Weisblum et al. 2020). Examination of almost 6500 B.1 genomes from the United States that were deposited in GISAID with collection dates between 2020-10-01 and 2021-02-26 revealed that only 4.4% harbored both S494P and P681H mutations. Of these, 68% derived from New York, representing the majority of cases (Supplementary Table 2). New Jersey, the state with the second highest number of B.1 variants had only 6% (Supplementary Table 2).

Since its initial detection in New York on 2020-12-01, the number of SARS-CoV-2 genomes with a P681H mutation attributable to this variant has risen, approaching 50% of all SARS-CoV-2 genomes with a P681H mutation in February 2021 (Supplementary Figure 1). The proportion of SARS-CoV-2 genomes containing P681H and S494P mutations in New York also increased by over 4% since December 2020 (Table 1 and Figure 4). This is less than the increase demonstrated by B.1.1.7 and both B.1.526 variants but greater than B.1.427 and B.1.429 (Table 1 and Figure 4), which carry the L452R mutation and were identified as rapidly spreading in California (Zhang et al. 2021; Tchesnokova et al. 2021). In fact, if we count the B.1 variants separately from the B.1 lineage, they ranked 12^th^ in abundance in January 2021 compared to other lineages detected and 5^th^ in February 2021, surpassed only by B.1 and B.1.2, which have been circulating in New York since the early days of the pandemic (Lasek-Nesselquist, Singh, et al. 2021; Maurano et al. 2020; Gonzalez-Reiche et al. 2020), and the B.1.1.7 and B.1.526 variants.

## Discussion

Multiple lineages have independently acquired a P681H mutation in New York and globally (Figure 2). Within New York, B.1.243 genomes with a P681H mutation represent ∼7% of the total number of SARS CoV-2 genomes sequenced. This variant showed an initial spike in frequency in late summer 2020 and slowly rose in prevalence only to decrease again in recent months. Sampling biases likely do not substantially influence the decline in the B.1.243 variant. While the regionally diverse sampling that occurred in September and October 2020 coincided with an uptick in COVID-19 cases due to this variant, it was already in decline by January 2021 when more New York counties were sampled at a greater depth (Figure 5). Analyses of B.1.243 in other states suggests that the rise of the B.1.243 variant in New York could be due to chance rather than increased fitness. Indeed, the initial spike of cases due to B.1.243 variants in October and November 2020 and a later smaller spike in December 2020 are associated with outbreaks in congregant settings, where founder effects could be responsible for rapid propagation.

**Figure 5.**
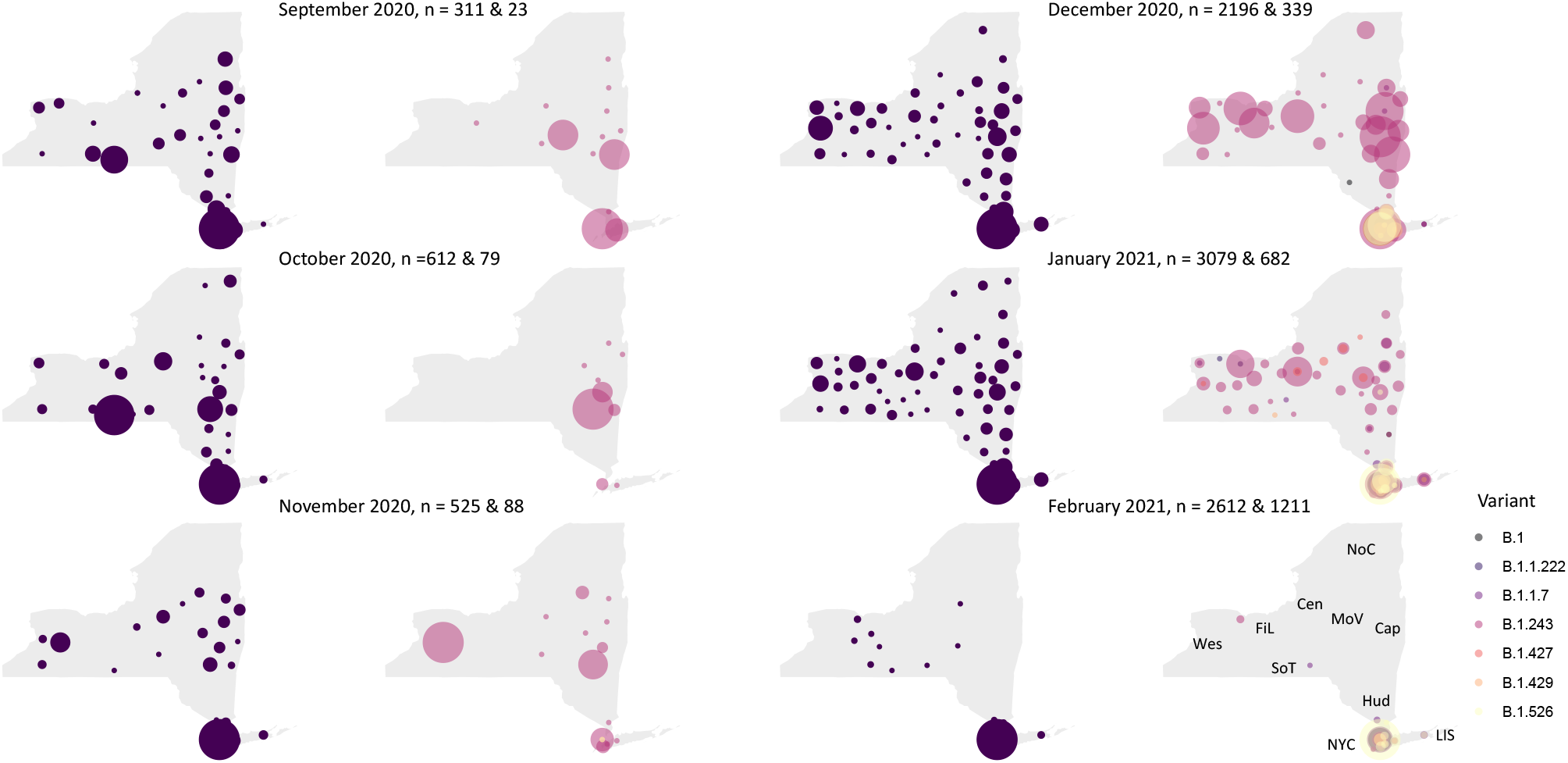
Bubble maps of SARS-CoV-2 sampling in New York. The total number of SARS CoV-2 (left-panel in pair) and variant genomes (right-panel in pair) mapped to their New York counties of origin since September 2020 (the first month which B.1.243 variants were detected). The number of SARS-CoV-2 genomes (n) in total (first number) and the number of variants (second number) sequenced per month are indicated. Bubble sizes are proportional to the number of genomes sequenced from each county. The ten regions of New York: Wes, Western; FiL, Finger Lakes; SoT, Southern Tier; Cen, Central; NoC, North Country; MoV, Mohawk Valley; Cap, Capital Region; Hud, Mid-Hudson; NYC, New York City; LIS, Long Island.

Alternatively, it is possible that P681H conferred a minor advantage to the B.1.243 lineage, allowing it to spread more easily in congregant settings, but was outcompeted by more recently emerged, highly transmissible variants, such as B.1.1.7 and B.1.526, and even other variants with a P681H mutation. Indeed, the dramatic decline of B.1.243 variants in February 2021 coincides with the explosion of other variants in New York (Figure 4). However, we cannot rule out the effects of some sampling bias as almost all SARS-CoV-2 genomes sequenced during this time period derived from the NYC metropolitan area (New York City, the Mid-Hudson, and Long Island), which had fewer B.1.243 variants in comparison to the rest of the state (Figure 5).

In contrast to the recent decline in B.1.243 variants, other variants with a P681H mutation carrying T478K or S494P mutations have modestly increased in frequency in New York. P681 is located five residues upstream of a multibasic site in the Spike protein, the cleavage of which facilitates processes (such as membrane fusion) required for viral entry into the cell (Shang et al. 2020; Papa et al. 2021). This mutation also occurs in B.1.1.7 and could contribute to its increased rate of transmission (Davies et al. 2021).

T478 is located in the RBD of the Spike protein, near the interface with ACE2 (Figure 6). A T478I mutation conferred moderate resistance to neutralization by two monoclonal antibodies and by convalescent serum from two patients (Liu et al. 2021). We would expect the T478K mutation to have a greater impact on neutralization, by introducing a larger residue with a positive charge at this position. The T478K mutation was identified in an in vitro evolution experiment to discover RBD mutations with increased affinity to ACE2, but it was not characterized further (Zahradnik et al., 2021). It is unclear why the T478K mutation would result in increased binding affinity, although it could be caused by either the change in the electrostatic potential in that region or by inducing a conformational change.

**Figure 6.**
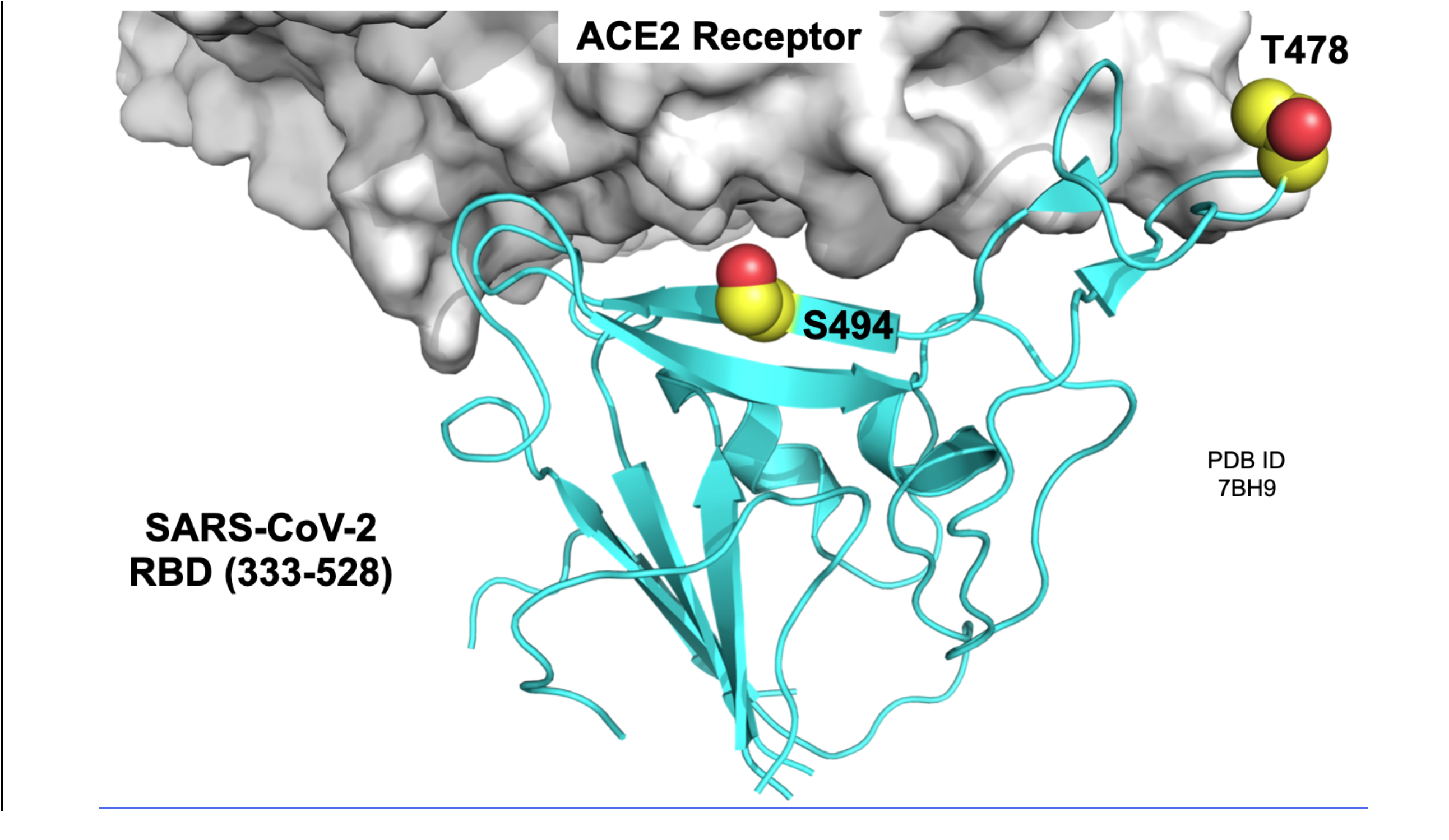
Sites of mutation at the interface between the SARS-CoV-2 spike protein RBD and the ACE2 receptor. Residues T478 and S494 are shown on the structure of the RBD bound to ACE2 (PDP ID 7BH9; Zahradník et al. 2021). The T478K mutation occurs together with P681H in the 20B variant and the S484P mutation occurs with P681H in the 20C variant. Figure 6 was prepared using the PyMOL Molecular Graphics System v2.3.5 (Schrödinger, LLC).

S494 is located in a beta strand in the RBD, at the interface with the ACE2 receptor (Figure 6). An in vitro screen for spike mutations that escape neutralization by monoclonal antibodies and convalescent serum identified S494P (Lui et al., 2021), which was later shown to reduce neutralization by 3-5-fold in some convalescent sera (Greaney et al. 2021b. However, this mutation was not as potent at neutralization as E484K (Greaney et al. 2021b) - a mutation harbored by B.1.526 variants circulating in New York.

While neither the B.1.1.222 nor the B.1 variant appears to be rising as dramatically as B.1.526 or B.1.1.7 within New York State, they have increased faster than the B.1.427 and B.1.429, which showed a rapid expansion in California (Zhang et al. 2021; Tchesnokova et al. 2021). However, we cannot rule out the effects of sampling biases, particularly for February 2021, when almost all specimens sequenced derived from the NYC metropolitan area - the location where the majority of B.1.1.222 and B.1 variants were circulating (Figure 5). Considering the phenotypic characteristics conferred by the mutations present in B.1 and B.1.1.222 variants, their moderate increase in New York, and that convergent evolution is often indicative of positive selection, we believe both variants are worth monitoring. However, if B.1.243 serves as an example, any potential selective advantage these variants have might be relatively inconsequential in the face of more transmissible or infectious forms, such as B.1.526 and B.1.1.7. Thus, this could be a case of enhanced surveillance noting variants of interest before they became a concern.

## Methodology RNA extraction

Respiratory swabs in viral transport medium (VTM, UTM or MTM) were received at the Virology Laboratory, Wadsworth Center, for SARS-CoV-2 diagnostic testing. Total nucleic acid was extracted using either easyMAG or eMAG (bioMerieux, Durham, NC), MagNA Pure 96 and Viral NA Large Volume Kit (Roche, Indianapolis, IN), or EZ1 with the DSP Virus Kit (QIAGEN, Hilden, Germany) following manufacturers’ recommendations. Extracted nucleic acid was tested for SARS-CoV-2 RNA using the 2019-Novel Coronavirus Real-Time RT-PCR Diagnostic Panel (Centers for Disease Control and Prevention) according to the Instructions for use on an ABI 7500Dx.

### Illumina library preparation and sequencing

Library preparation and whole genome amplicon sequencing of SARS-CoV-2 was performed using a modified ARTIC protocol (https://artic.network/ncov-2019) as described previously (Lasek-Nesselquist, Lapierre, et al. 2021).

### Bioinformatics processing

Reads were processed with ARTIC nextflow pipeline (https://github.com/connor-lab/ncov2019-artic-nf/tree/illumina, last updated April 2020) as previously described (Lasek-Nesselquist, Lapierre, et al. 2021).

### Analysis of New York SARS-CoV-2 genomes and detection of emerging variants

All SARS-CoV-2 genomes from New York were downloaded from GISAID as of 2021-03-04 or 2021-02-26. Genomes were annotated with NextClade (https://clades.nextstrain.org/), the results of which were inspected for changing trends in the data and further queried for mutations of interest. Pangolin lineages, geographic locations, and collection dates for genomes carrying mutations of interest were then extracted from the associated metadata provided by GISAID. Lineage frequencies were calculated on a monthly basis by dividing the number of genomes of a lineage by the total number of genomes sequenced each month. This was also performed for mutations and variants of interest. Stacked area plots and bubble maps were rendered in R v4.0.3 (https://www.r-project.org/) with ggplot2 and map packages and coloring provided by the viridis package.

### Phylogeographic analysis

We constructed a time-resolved phylogeny of SARS-CoV-2 genomes in Nexstrain v2.0.0 (Hadfield et al. 2018) using representatives of B.1.243 and B.1.1.22 lineages and contextual sequences sampled from New York and the rest of the world (see Acknowledgements table for genomes used and contributing laboratories). In summary, a total of 2689 genomes >27 Kb in length were aligned in MAFFT v7.475 (Katoh & Standley 2013), a maximum likelihood phylogeny was generated with IQ-TREE v2.0.3 (Nguyen et al. 2015) under a GTR+G4 nucleotide substitution model, and divergence times as well as ancestral traits were inferred with TreeTime (Sagulenko et al. 2018). The clock rate was fixed at 8×10^−4^ substitutions per site per year. Trees were visualized in Auspice (Hadfield et al. 2018) and with the ggtree (Yu et al. 2017) package for R. The timing of introduction for the B.1 variant containing P681H and S494P mutations was estimated from a previous phylogeographic analysis (Lasek-Nesselquist, Lapierre, et al. 2021).

### IRB Approval

This work was approved by the New York State Department of Health Institutional Review Board, under study numbers 02-054 and 07-022.

## Supporting information

Acknowledgement table

## Data Availability

All SARS CoV-2 genomes are available on GISAID.org

## Acknowledgements

Next generation sequencing was performed by the Advanced Genomic Technologies Core of the Wadsworth Center. Initial funding for sequencing was generously provided by the New York Community Trust. This publication was also supported by Cooperative Agreement number NU50CK000516, funded by the Centers for Disease Control and Prevention. Its contents are solely the responsibility of the authors and do not necessarily represent the official views of the Centers for Disease Control and Prevention or the Department of Health. We graciously thank all originating and submitting laboratories for their SARS-CoV-2 sequence contributions to the GISAID database and Wadsworth Center’s Bioinformatics Core. We also humbly thank Paul Masters for his suggestions.

## Author contributions

Conception, data analyses, and writing of the manuscript were performed by ELN. Figure 6 was generated and structural analysis of T478K and S494P mutations were conducted by JP. Manuscript revisions were performed by ELN, JP, ES, and KSG.

---

**Supplementary Table 1.**
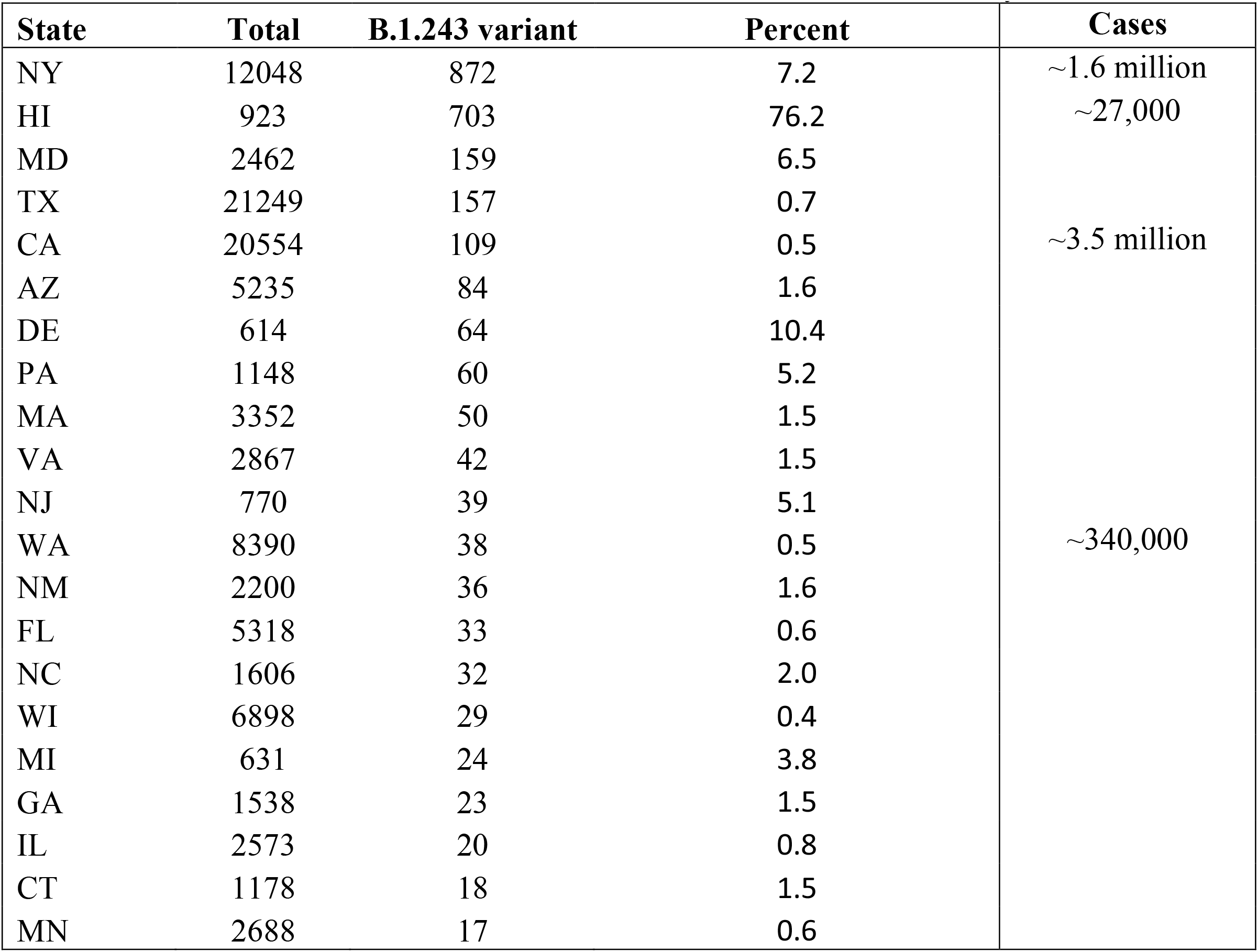
The Number of B.1.243 SARS-CoV-2 genomes with P681H mutations for select states. Total; the total number of genomes sequenced as of 2021-02-26 for states with the greates number of B.1.243 variants; Percent, the proportion of the total number of SARS-CoV-2 genomes that are a B.1.243 variant; Cases, the total number of COVID-19 cases for select states. Number of cases was taken from www.nytimes.com

**Supplementary Table 2.** States that have sequenced B.1.1.222 SARS-CoV-2 genomes and the number that contain spike mutations T478K and P681H. Total number of B.1.1.222 genomes identified (as of 2021-02-26) in 48/50 states and the proportion that are variants.

| State | T478K & P681H | Total | % Variant |
| --- | --- | --- | --- |
| TX | 145 | 260 | 56 |
| CA | 133 | 438 | 30 |
| **NY** | **82** | **125** | **66** |
| WA | 46 | 73 | 63 |
| IL | 33 | 53 | 62 |
| LA | 33 | 38 | 87 |
| MD | 33 | 64 | 52 |
| FL | 32 | 56 | 57 |
| MO | 27 | 27 | 100 |
| MI | 26 | 43 | 60 |
| KS | 21 | 30 | 70 |
| CO | 20 | 109 | 18 |
| IN | 11 | 13 | 85 |
| NV | 11 | 13 | 85 |
| NM | 9 | 42 | 21 |
| UT | 9 | 16 | 56 |
| WI | 9 | 61 | 15 |
| OR | 8 | 45 | 18 |
| AK | 7 | 8 | 88 |
| GA | 6 | 12 | 50 |
| MA | 6 | 8 | 75 |
| OH | 6 | 6 | 100 |
| WY | 6 | 28 | 21 |
| AL | 5 | 8 | 63 |
| KY | 5 | 5 | 100 |
| MN | 5 | 16 | 31 |
| NJ | 5 | 11 | 45 |
| PA | 5 | 7 | 71 |
| AZ | 4 | 15 | 27 |
| NC | 4 | 12 | 33 |
| AR | 3 | 3 | 100 |
| OK | 3 | 3 | 100 |
| DC | 2 | 2 | 100 |
| NH | 2 | 2 | 100 |
| PR | 2 | 3 | 67 |
| VA | 2 | 8 | 25 |
| DE | 1 | 2 | 50 |
| HI | 1 | 1 | 100 |
| IA | 1 | 3 | 33 |
| ID | 1 | 1 | 100 |
| MS | 1 | 1 | 100 |
| MT | 1 | 2 | 50 |
| NE | 1 | 6 | 17 |
| RI | 1 | 1 | 100 |
| CT | 0 | 1 | 0 |
| ME | 0 | 1 | 0 |
| SC | 0 | 3 | 0 |
| TN | 0 | 2 | 0 |

**Supplementary Table 3.** The number of B.1 variants (those with S494P and P681H mutations) for states with at least one COVID-19 related case. Percent is the proportion of the total number of variants identified (283) as of 2021-02-26.

| **State** | **S494P & P681H** | **Percent** |
| --- | --- | --- |
| NY | 193 | 68.2% |
| NJ | 18 | 6.4% |
| MI | 16 | 5.7% |
| MA | 8 | 2.8% |
| MD | 7 | 2.5% |
| TX | 7 | 2.5% |
| MN | 5 | 1.8% |
| PA | 4 | 1.4% |
| WI | 4 | 1.4% |
| DE | 3 | 1.1% |
| CA | 2 | 0.7% |
| ME | 2 | 0.7% |
| PR | 2 | 0.7% |
| CO | 1 | 0.4% |
| CT | 1 | 0.4% |
| FL | 1 | 0.4% |
| GA | 1 | 0.4% |
| IA | 1 | 0.4% |
| LA | 1 | 0.4% |
| NC | 1 | 0.4% |
| RI | 1 | 0.4% |
| VT | 1 | 0.4% |
| WA | 1 | 0.4% |
| WV | 1 | 0.4% |
| Unknown | 1 | 0.4% |

**Supplementary Figure 1.**
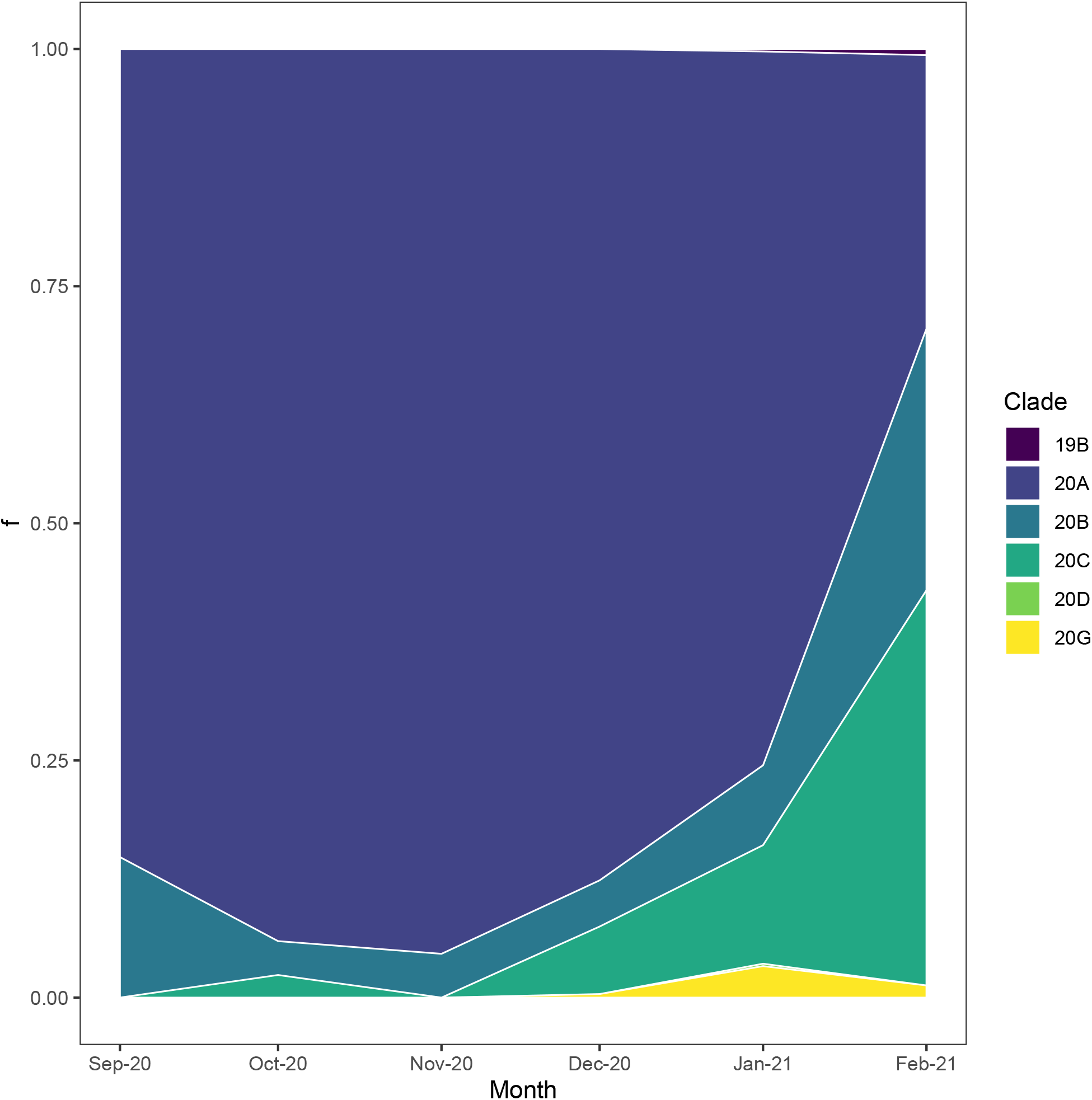
Proportion of genomes in major Nextstrain clades that contain a P681H mutation in New York State. Stacked area plot of the proportion of Nextstrain clades containing P681H mutations in New York State through time. 20A mainly represents the B.1.243 lineage and 20C mainly represents B.1 variants with P681H and S494P mutations. 20B represents the frequency of both B.1.1.7 and B.1.1.222 lineages.

